# Social media language patterns reveal early signs of impending stroke: An observational study

**DOI:** 10.1101/2025.11.05.25339635

**Authors:** Yael Dahari, Tal Ariel Ziv, Elad Yom-Tov

## Abstract

**Background:** Stroke remains a major global health concern, contributing substantially to mortality and long-term disability. Current clinical tools lack effective mechanisms for early detection. Here, we investigate whether linguistic and behavioral patterns in online social media can serve as early indicators of impending stroke.

**Methods:** We analyzed posts from 1,683 Reddit users who reported experiencing a stroke and 2,438 users across three control groups al made between 2015 and 2024. Linguistic features were extracted from posts, and predictive models were trained to distinguish individuals who experienced a stroke from controls.

**Results:** Our results reveal changes in several linguistic markers, e.g., the rate of spelling errors and the use of word tokens, beginning approximately 20 weeks prior to the stroke. Using posts from the four months preceding the event, the predictive model achieved an area under the curve (AUC) of 0.87.

**Discussion:** These findings highlight the potential of social media–derived linguistic signals to predict strokes several months in advance, offering a promising avenue for early detection and preventive interventions in digital medicine.

## Introduction

Stroke is the second leading cause of death worldwide and a major contributor to long-term disability (1)}. Its incidence continues to rise, particularly among younger populations in low- and middle-income countries. Preventing stroke is considerably more effective than treating it after onset. About 87% of strokes are considered preventable through management of modifiable risk factors, including hypertension, hyperlipidemia, obesity, and hyperglycemia (2).

Current diagnostic methods for stroke rely on physical examination, blood tests, and neuroimaging to confirm stroke type and assess brain damage (3). While these approaches are highly effective in symptomatic individuals who are suspected of having a stroke, they are impractical for large-scale screening because of their cost and limited accessibility (4).

At present, stroke screening typically uses cardiovascular risk scoring systems such as the American Heart Association’s cardiovascular disease risk algorithm or the Framingham risk score. However, these tools estimate the likelihood of any cardiovascular event, not stroke specifically. Moreover, they assess the risk of an event over a ten-year period rather than estimating short-term stroke risk.

We hypothesize that one of the problems in screening for stroke is the subtlety of symptoms in comparison to available data. Specifically, the changes in people’s behavior need to be assessed over a period of time, but clinical data are often sampled at intervals that are too large to allow such estimation. In this context, internet data, including interactions with search engines, posts on social media, and chats with Large Language Models (LLMs) may offer a viable data source for stroke detection.

Recently, Shaklai et al. (5) showed that changes in people’s behavior observed while they interact with search engines can be used to differentiate people at risk of stroke from healthy controls. Furthermore, these changes are predictive of stroke up to 4 months prior to the event. However, search engine queries are short, and so we hypothesize that using richer text may result in better predictive ability.

Thus, here we analyze social media posts to identify patterns indicative of stroke risk. Social media platforms have been used for early detection of other health conditions. For example, Reddit discussions have been shown to predict transition to suicidal ideation (6,7), anxiety (8) and depression (9,10). However, few studies have used social media to predict physical conditions and did so using the words in the posts, not attributes derived from them. For example, Eichstaedt et al. (11) predicted county-level heart disease mortality from Twitter data. Merchant et al. (12) examined the additional predictive value of Facebook posts over demographics to predict 21 medical conditions.

Reddit is an especially suitable platform for such research because of its unique characteristics which include anonymity, a large number of users, and rich content. First, users are more likely to discuss sensitive or stigmatized topics openly when anonymous (13). With over 108 million daily active users, Reddit contains billions of posts (14), enabling the construction of large cohorts. Finally, unlike platforms with strict character limits (e.g., Twitter), Reddit allows long-form posts (up to 40,000 characters), providing nuanced insights into user behavior and language patterns.

In this paper, we aim to ascertain whether changes in language can be observed prior to stroke, to develop a method for detecting individuals at high risk of stroke with sufficient lead time to enable interventions to prevent stroke.

## Methods

The research described herein was approved by the Ethics Committee of the Faculty of Exact Sciences at Bar Ilan University on January 2023 (no specific approval number was given). The research was conducted in accordance with the Declaration of Helsinki.

### Data

We extracted all Reddit posts from 2015 to 2024 (inclusive) that indicated that the writer, or someone close to them, may have experienced a stroke. Specifically, we searched for users who used one of the following phrases: self (“I have”, “I was diagnosed with”, “I had”, “I experienced”) and others (“my wife”, “my husband”, “my spouse”, “my boyfriend”, “my girlfriend”, “my partner”) followed (within 20 characters) by one of the phrases: “stroke”, “cerebrovascular accident”, “CVA”, “cerebrovascular event”, “transient ischemic attack”, “brain infarction”, “brain ischemia”, “cerebral ischemia”.

We then extracted all the posts of users who matched these phrases (from Reddit in its entirety) and kept users with at least 5 posts (to ensure sufficient data per user) and no more than 5000 posts during the data period (to reduce the likelihood of including automated accounts).

### Data labelling

We used a Large Language Model (LLM) ChatGPT-4o to label each post for several key attributes related to stroke context, including relevance, the relationship between the author and the stroke patient, demographics (age and gender), and attributes of the stroke event. The full list of annotated attributes is shown in Table 1.

**Table 1:** List of Annotated Attributes.

| Num | Attribute | Possible values | Comments |
| --- | --- | --- | --- |
| 1 | Stroke Relevance | 0 (Unknown), 1–5 | Ordinal: 1 = Not relevant, 5 = Definitely stroke-related |
| 2 | Who Had the Stroke | Author, Sibling, Partner, Parent, Friend, Other, Unknown | Relationship between post author and the person who experienced the stroke |
| 3 | Age group of the person | 17 or less, 18–25, 26–44, 45–64, 65–74, 75–84, 85+, Unknown | Age group of the stroke patient |
| 4 | Gender of the person | Male, Female, Other, Unknown |  |
| 5 | Date of stroke | Exact date (DD.MM.YYYY), or relative date (e.g., ``two weeks ago''), or unknown | -1 denoted unknown |
| 6 | Certainty of stroke | Yes, No, Unknown | Indicates whether a stroke diagnosis was confirmed |
| 7 | Is this the first stroke? | First stroke, Second or more, Unknown | If not mentioned, assumed to be the first stroke |
| 8 | Cause of stroke | Mental stress, Drug use, Illness, Other, Unknown | Based on explicit mention in the post |

To validate our labelling protocol, two of the authors independently labelled 50 randomly selected posts. Inter-annotator agreement was measured using Spearman’s rank correlation for ordinal variables and Cohen’s Kappa for categorical variables. We also labelled the same 50 posts using ChatGPT-4o and compared its labels to the human annotations using the same metrics. Given that sufficient agreement was observed between human annotators and between humans and the LLM, we used the LLM to label the full dataset.

### Thresholding and Filtering

Low relevance (attribute 1) scores rendered other answers inapplicable. Therefore, we first used ChatGPT to tag posts for relevance and filtered out low-relevance posts. We then manually labeled another 50 posts for the remaining questions.

To determine appropriate cutoff values for ordinal scores, we converted one set of labels to binary values at various thresholds and compared those with the other labeler. We did this to compare the two authors and then again between one of the authors and the model.

### User Groups

We divided our users into four groups:

- **Timed Stroke Group**: Users who reported experiencing a stroke and where the stroke was dated to within a month or less.
- **Control Group 1**: Users who experienced a stroke, but for whom we did not know the date of the stroke with sufficient accuracy.
- **Control Group 2**: Users who are related to someone who experienced a stroke (e.g., siblings, spouses, friends), but did not experience one themselves.
- **Control Group 3**: Users who mentioned a stroke in their posts, but were not members of the previous groups (that is, neither they or someone close to them experienced a stroke).

### Extracting Post-Level Attributes

We extracted from each Reddit post authored by users in our dataset a set of attributes potentially reflecting linguistic and cognitive functioning. These attributes included measures of lexical diversity, readability, and syntactic complexity, with the full list provided in Table 2. For each user, we computed summary statistics for these attributes—including average, median, standard deviation, maximum, minimum, and temporal slope—across the study period. We further incorporated available demographic information (age and gender) from the tagged posts, as well as each user’s posting frequency during the observation window.

**Table 2:** Linguistic, Readability, Cognitive, and Cohesive Metrics extracted from user texts.

| Metric | Range in dataset | Comments |
| --- | --- | --- |
| Average number of words per post | 0 -- 1,000 |  |
| Average word length | 0 -- 50 |  |
| Average number of words per sentence | 0 – 125 |  |
| Fraction of unique words in a post | 0.0 – 1.0 |  |
| Syntactic complexity | 0 -- 5 |  |
| Flesch-Kincaid readability score (15) | 0 -- 50 |  |
| Dale-Chall readability score (16) | 0 – 25 |  |
| Moving average Type–Token Ratio (MATTR) (17) | 0.4 – 1.0 | Lexical diversity over a sliding window |
| Root Type–Token Ratio (TTR) (17) | 0.0 – 17.5 | Lexical diversity normalized by root of token count |
| Fraction of spelling errors | 0.0 – 1.0 |  |
| Cohesion score (18) | 0 – 12.5 |  |
| Average POS ratio (18) | 0.0 – 1.0 | The ratio of distinct part-of-speech tags to total tokens per sentence, averaged |
| POS variation (verb/noun/adj) (18) | 0.0 – 1.0 | How many unique verb, nouns and adjective lemmas are used relative to total |
| Sentiment score | -1.0 – 1.0 |  |
| Existence of stroke-related keywords | True / False | Mentions of stroke-related terms like cardiovascular conditions, blood pressure, migraines, blood thinner. The full list is shown in the Appendix. |
| Average word probability | 0 -- 0.03 | Based on Google NGRAM frequency (19) |

To align features in time, posts were indexed relative to either the stroke event (as explicitly reported in posts from the timed stroke group) or, for control groups, the midpoint between each user’s first and last post.

### Predictive modelling

We trained machine learning models on the independent attributes described above to predict whether a user belonged to the timed stroke group or to each of the control groups. Models were evaluated across multiple temporal windows prior to the stroke event: 3, 6, 12, and 24 months. The 3-month window yielded the highest AUC scores and was therefore selected as the primary analysis window.

We experimented with several classifiers, including Random Forest, Logistic Regression, and gradient boosting. Results reported here are based on gradient boosting with the XGBoost algorithm, which achieved the best predictive performance.

Model performance was assessed using the area under the ROC curve (AUC). To mitigate overfitting, we applied 5-fold cross-validation in all experiments.

## Results

### Population statistics

A total of 27,120 posts from 11,990 users mentioned stroke. After filtering for relevance (Item #1) and applying thresholds on posting activity, the dataset was reduced to 4,121 users. These were distributed as follows: 1,683 users in the timed stroke group, 1,319 in Control Group 1, 523 in Control Group 2, and 596 in Control Group 3.

In the timed stroke group, slightly more users identified as female than male (53% vs. 47%). A similar distribution was observed in Control Groups 1 (53%) and 3 (56%), whereas Control Group 2 had more males (58%). The most frequently reported age range was 26–44 years (see Appendix for the full distribution). These demographic patterns are due to both stroke incidence and the age distribution of Reddit users.

When the number of stroke events was mentioned, over 93% of users reported a first stroke, with the remainder reporting a second or subsequent event. Control Group 3 differed slightly, with 87% first strokes. Illness was the most commonly cited cause of stroke (45% in the timed stroke group); a full breakdown is provided in the Appendix.

Posting frequency did not differ significantly across groups (Kruskal–Wallis test: H = 4.38, p = 0.22).

Agreement between human annotators was high (rho > 0.89, kappa >= 0.65). Agreement between humans and the LLM was slightly lower but still strong (rho > 0.82, kappa > 0.53). Full results are shown in the Appendix. These findings suggest that the LLM labels posts in a manner closely aligned with human annotations.

### Textual attributes over time

We computed average weekly values of each attribute relative to each user’s stroke date (or other relevant date, as described above). Figure 1 shows three measures of interest over time: the Type– Token Ratio normalized by the square root of the token count (MATTR), the average ratio of unique words per post, and the average fraction of spelling mistakes per post. Additional variables are shown in the Appendix.

**Figure 1:**
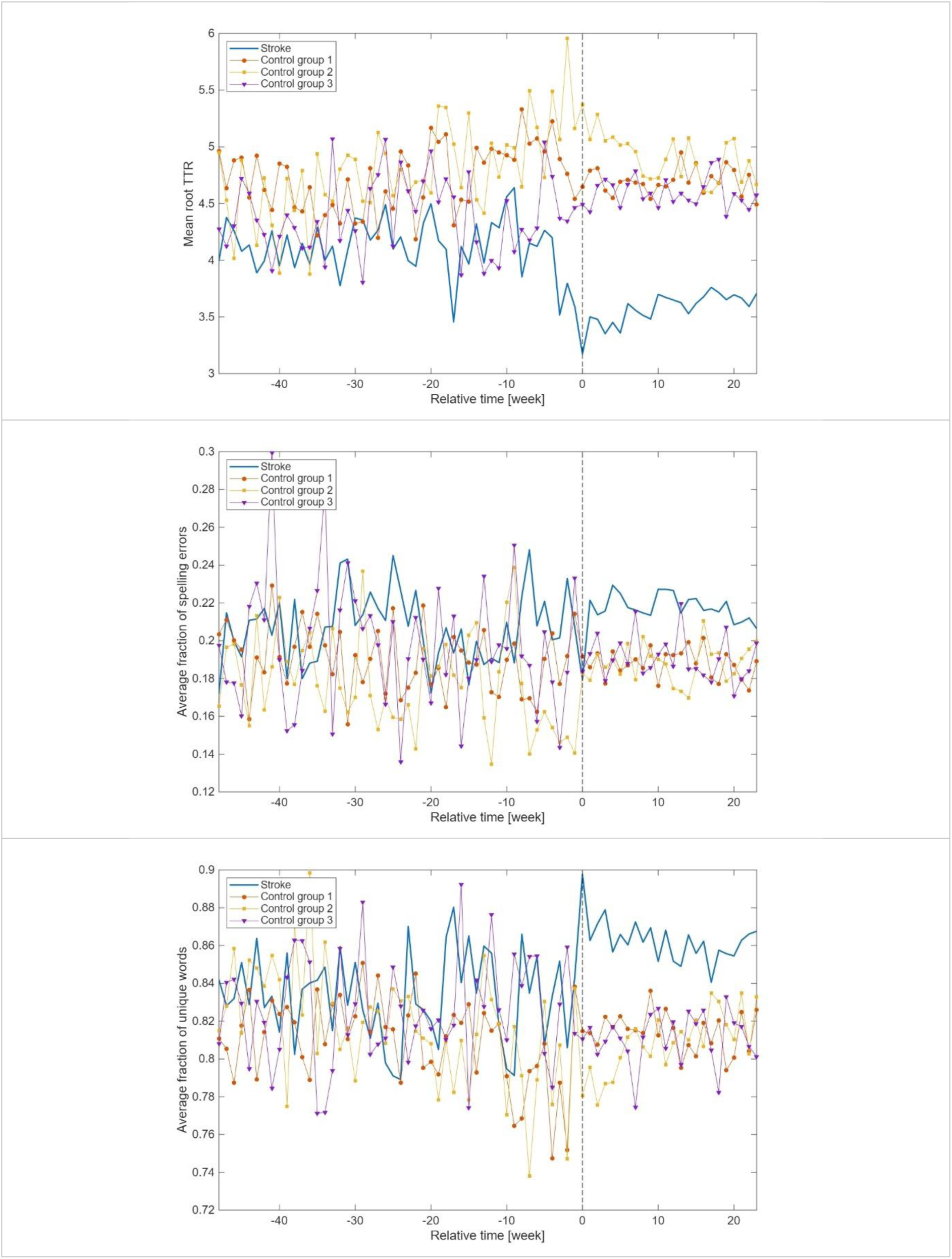
Measures of user texts over time, stratified by user groups: Weekly Mean Root TTR (top), average fraction of unique words (center) and fraction of spelling errors (bottom).

First, as the figure shows, the MATTR of all groups begins with similar values, but around 16 weeks before the stroke, the timed stroke group begins to decline. This trend accelerates immediately prior to the stroke and though it rises slowly, it remains significantly below the baseline for at least 25 weeks post-stroke.

All groups begin with a similar average fraction of unique words. The timed stroke group (and to some extent Control Group 3) show a notable increase in values from around 20 weeks prior to the stroke. Although a higher unique word fraction might suggest a richer vocabulary use by the authors, we hypothesized that this increase may be due to spelling errors. To test this, we examined the fraction of spelling mistakes in the text and observed a similar trend whereby all groups start at similar values, but the timed stroke group shows a clear upward trend in the fraction of spelling errors, especially post-stroke.

### Modelling the risk of stroke

We evaluated model performance over time using a sliding 3-month window starting two years prior to the stroke. This analysis was conducted separately for distinguishing members of the timed stroke group from each of the control groups.

Figure 2 shows the AUC over time. The model most easily distinguishes the timed stroke group from Control Group 2 (relatives of people who experienced a stroke), whereas the other two groups are more difficult to differentiate from the timed stroke group.

**Figure 2:**
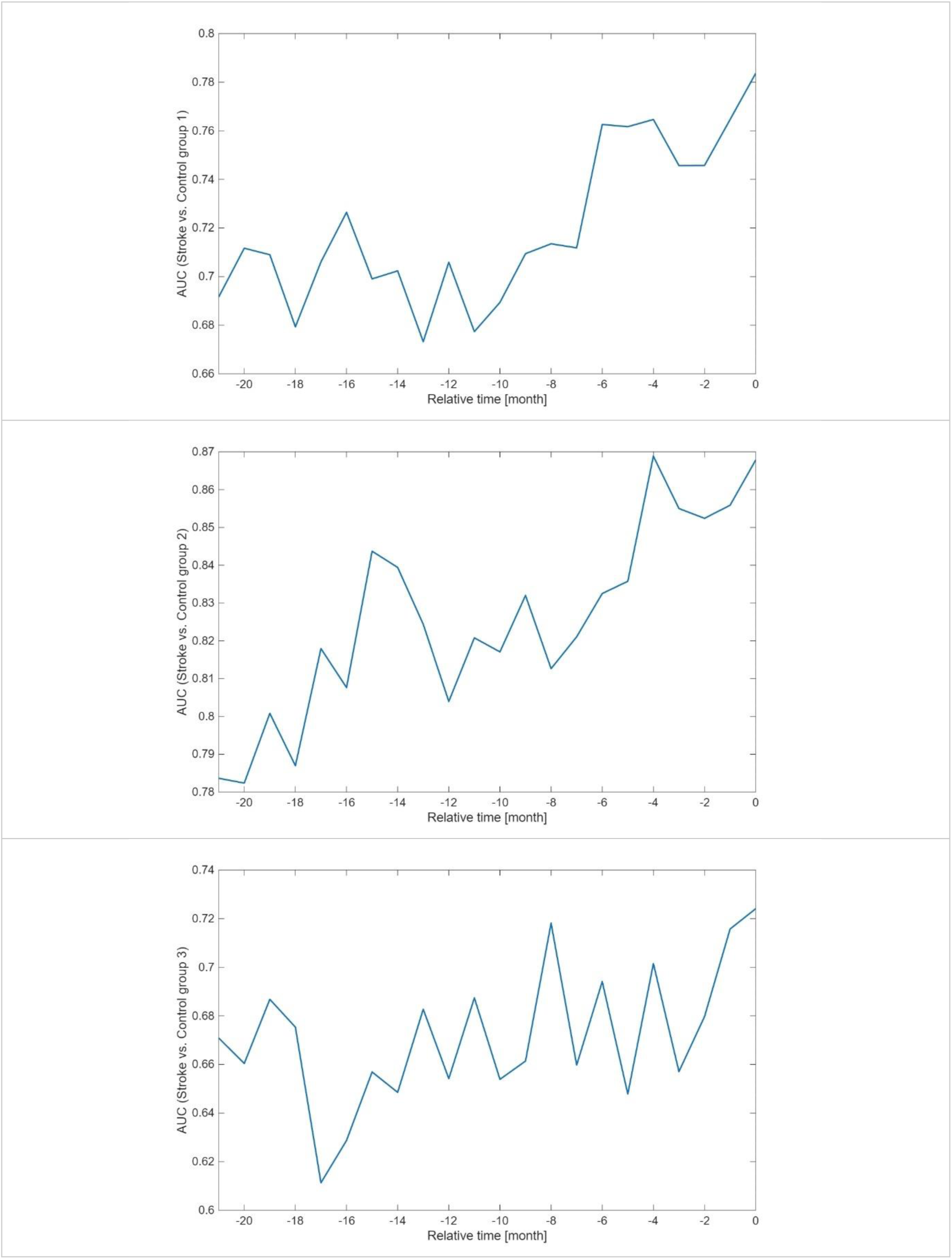
AUC over time when differentiating the timed stroke group from control groups 1 (top) to 3 (bottom). Note the different values of the vertical axis.

Model performance for Control Group 3 remains nearly constant over time. In contrast, when distinguishing the timed stroke group from Control Group 1 (users with an untimed stroke) and, to a lesser extent, Control Group 2, performance is relatively stable until approximately 10 months prior to the stroke. At that point, the AUC begins to rise—for Control Group 1, from roughly 0.68 to 0.78. This pattern suggests that changes in the timed stroke group around 10 months before the stroke make these users increasingly dissimilar from the control groups.

We evaluated the additional predictive value of age and gender by training models with and without these demographic variables for each control group across 3-, 6-, 12-, and 24-month time windows. The only statistically significant improvement in AUC (20) was observed when distinguishing the timed stroke group from Control Group 2 (P < 10^-6^ for all time windows). Thus, demographic information primarily contributes to differentiating individuals who experienced a timed stroke from their relatives.

Finally, we analyzed model prediction scores of the timed stroke group stratified by attributed cause. Scores differed significantly overall (Kruskal–Wallis test: H = 78.8, P < 0.0001), but post-hoc pairwise comparisons (Bonferroni-corrected, P < 0.05) revealed significant differences only between individuals reporting stroke due to drug use, illness, or mental stress versus those with unknown causes. In all three cases, scores were lower than those of the unknown-cause subgroup, indicating that strokes with an attributable cause were more easily detected than strokes of unknown cause.

## Discussion

Stroke is a leading cause of disability and mortality, yet no methods exist to effectively screen individuals for it. Recently, it has been suggested that changes in language use in internet search engines predict stroke (5). Here we analyzed Reddit posts made over a 10 year period and compared users who indicated that the author had experienced a stroke to users in three control groups. We examined linguistic and cognitive features extracted from Reddit posts over time, alongside demographic metadata where available. We also trained Machine Learning classifiers to distinguish each control group from the timed stroke group based on features in the period leading up to the stroke.

We identified 1,683 Reddit users who experienced a stroke and for whom that stroke could be accurately timed. Users who experienced a stroke (and indicated their demographics) were slightly more likely to be female and young. The literature shows that stroke is more common among older individuals and males (21), but our findings represent the demographics of Reddit users which skews towards younger ages. Over 93% of cases in which the number of stroke event was known, it was the first stroke. This is in contrast with past findings, which showed that approximately 25% of hospitalizations for stroke were recurrent stroke events (22). We attribute this to people’s tendency to post on new events, rather than recurring events (23).

When examining feature trajectories over time, we observed that all groups were similar to each other one year before the stroke date. However, beginning approximately 16 weeks prior to the event and continuing through the six-month post-stroke period, the timed stroke group diverged from the controls in several linguistic and cognitive measures.

For instance, the weekly Mean Root TTR decreased in the timed stroke group after the stroke compared to before, while no significant change was observed in the control groups. Similarly, the weekly mean Unique Word Fraction displayed a clear distinction between groups: after the stroke, the timed stroke groups’ values increased, whereas the control groups maintained consistent levels. This observation aligns with prior findings that lexical diversity measures such as MATTR effectively differentiate individuals with aphasia from neurotypical controls and those with mild aphasia (24).

Another feature that differed markedly was the spelling error rate. This feature spiked in the timed stroke group approximately eight weeks before the stroke and remained elevated throughout the six-month post-stroke period. This finding is supported by research indicating that agraphia, an acquired neurological disorder characterized by impaired writing ability despite previously normal skills, can be an early symptom of stroke (25).

Finally, we observed changes in the Unique Word Fraction per post. Approximately 20 weeks before the stroke, this measure began to rise, and post-stroke, the timed stroke group exhibited the highest values. Although previous research reported that stroke patients typically produce significantly fewer infrequent words compared to healthy controls, implying a decrease in lexical diversity (26), the observed increase in our study may be explained by the elevated spelling error rate. Misspelled words are treated as distinct tokens, which artificially inflates lexical diversity measures.

Our analysis found that demographic variables and average posting frequency were generally similar across groups, suggesting that predictive ability was not based on superficial features such as demographics or activity level.

The predictive model performed particularly well in distinguishing the timed stroke group from Control Group 2 (users related to someone who experienced a stroke). Interestingly, the separability between these groups was evident even two years prior to the stroke event. This may be an indication of quantifiable risk factors in the stroke population. Moreover, model performance improved as the stroke date approached, suggesting the emergence of pre-stroke changes in language use over time. The model had lower performance when distinguishing the timed stroke from Control Group 1 (users who had a stroke, but its timing is unknown). We attribute this to the fact that both groups were at-risk for stroke (e.g., age). However, model performance improved markedly around 10 months before the stroke date of the timed stroke group (AUC: 0.78). This is expected, as timed stroke users were approaching their actual stroke date, while users in the control group were aligned to a random time point.

Finally, model performance for Control Group 3 remained relatively low (AUC over time between 0.65 and 0.69) and exhibited no clear temporal trend. This group included users who mentioned a stroke but were classified as not having had one. Some of these users may have been mislabelled, or may share similar behavioral patterns with the timed stroke group due to other medical or psychological conditions, complicating the model’s ability to distinguish them reliably.

Taken together, these results suggest that linguistic and cognitive features not only distinguish stroke-affected individuals from non-affected controls a long time prior to the stroke event, but become more pronounced as the actual stroke event approaches. The difficulty in differentiating the timed stroke group from Control Group 1 highlights that both groups share a similar overall risk profile, but the temporal strengthening of model performance underscores that pre-stroke linguistic signals intensify closer to the stroke date. This temporal pattern supports the hypothesis that certain language and cognitive markers could serve as early indicators of stroke vulnerability, with increasing predictive power as the event nears.

Our work has several limitations. First, a central component of our methodology relied on the ability of a Large Language Model (ChatGPT 4o) to label users and posts based on their content. As with any automated system, misclassifications are possible. However, we verified that the model extracts labels in agreement levels similar to those observed between humans.

Another limitation involves missing data. Several features, particularly demographic ones, were labelled as “unknown” for a large proportion of cases. For instance, gender information was unavailable for 69% of users. This lack of complete metadata likely impaired the model’s ability to leverage potentially informative attributes and may have introduced bias.

Additionally, the reliability of self-reported data on Reddit remains a concern. We cannot verify the authenticity of users’ claims about having had a stroke, age, gender, or timing. Some posts may be exaggerated, fabricated, or produced by non-human accounts despite our filtering.

Despite these limitations, our findings provide early evidence that subtle linguistic changes detectable in social media writing may precede clinical stroke events, potentially enabling pre-emptive health interventions.

## Author contributions

EYT initiated the research, helped analyze the data, and supervised the project. YD and TAZ collected the data and analyzed it.

All authors wrote, read and approved the final manuscript.

## Data availability

Reddit data is publicly available, for example, via Academic Torrents. All labels generated for this work and other processed data is available from the authors upon reasonable request.

## Code availability

The code will be made publicly available on Github once the paper is accepted.

## Clinical trial number

Not applicable

## Appendix

### Appendix 1: The Full List of Stroke-Related Keywords

We looked for mentions of the following stroke-related terms:

1. **Headache and migraine terms**: headache, migraine, pain, symptom, drug, seizure.
2. **Stroke and neurological terms**: stroke, cerebrovascular, cerebral infraction, cva.
3. **Cardiovascular conditions**: heart attack, myocardial infraction, blood pressure, hypertension, clot, embolism, thrombosis, angina, cardiovascular.
4. **Blood pressure medication**: lisinopril, amlodipine, losartan, hydrochlorothiazide, metoprolol, atenolol, norvasc, cozaar, zestril.
5. **Anticoagulants / Blood thinners**: warfarin, coumadin, xarelto, eliquis, plavix, aspirin, clopidogrel, heparin, blood thinner, anticoagulant.

#### Attribution of stroke events

Table 3 shows the attribution of stroke events. As indicated in the table, in the vast majority of cases, users self-reported their own stroke experiences. Only a small fraction of posts referred to stroke events in others, such as partners, parents, or friends.

**Table 3:** Stoke Occurrence Among Users.

| Option | Count | Percentage |
| --- | --- | --- |
| Author | 10442 | 87 |
| Partner | 1177 | 10 |
| Other | 132 | 1 |
| Parent | 102 | 1 |
| Unknown | 80 | <1 |
| Friend | 40 | <1 |
| Sibling | 9 | <1 |

#### Age distribution

Table 4 shows the age distribution of users who reported having a stroke, based on inferred or declared age. The majority of stroke events occur in users aged 26–44, followed by the 18–25 and 45–64 age groups. Reports among older age groups (65+) are rare, which may reflect platform demographics, as younger adults are more likely to use Reddit. The low number of reports among those under 18 also supports the assumption that strokes in this age group are rare.

**Table 4:** Age Distribution in Percentage By Group.

| Group | 17 or less | 18-25 | 26-44 | 45-64 | 65-74 | 75-84 | 85 plus |
| --- | --- | --- | --- | --- | --- | --- | --- |
| Timed stroke | 1 | 32 | 56 | 9 | 1 | <1 | <1 |
| Control group 1 | 12 | 31 | 47 | 9 | 1 | <1 | <1 |
| Control group 2 | 3 | 10 | 44 | 32 | 7 | 3 | 1 |
| Control group 3 | 5 | 51 | 39 | 5 | <1 | <1 | <1 |

#### Stated cause of stroke

The stated cause of the stroke is shown in Table 5. As the table shows, the most commonly attributed cause for the stroke was illness (except for control group 3).

**Table 5:**
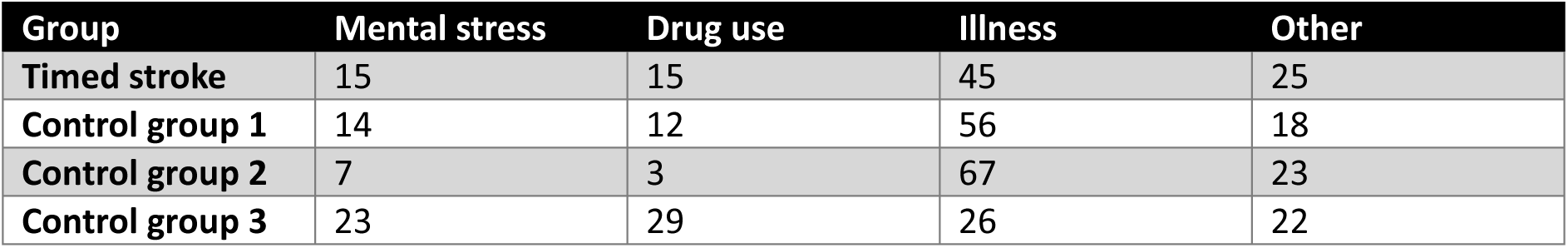
Attributed cause of the stroke in each group (percent)

#### Agreements on Labels

Table 6 shows the inter-human agreement and the human-LLM agreement for the different questions.

**Table 6:** Annotator agreement. The measures of agreement are Spearman rho for ordinal variables and Cohen’s Kappa for categorical variables. All results are statistically significant at P<0.0001.

| Question num | Measure | Inter-human | Human-LLM |
| --- | --- | --- | --- |
| 1 | Rho | 0.89 | 0.82 |
| 2 | Kappa | 0.90 | 0.74 |
| 3 | Rho | 0.90 | 0.83 |
| 4 | Kappa | 0.75 | 0.58 |
| 6 | Kappa | 0.80 | 0.71 |
| 7 | Kappa | 0.65 | 0.53 |
| 8 | Kappa | 0.78 | 0.62 |

The results indicate high agreement among human annotators across both ordinal and categorical questions. The agreement between humans and the LLM is slightly lower, though still substantial for most questions, especially for the ordinal ones. While agreement is lower for some categorical questions (e.g., Q7), the scores remain statistically significant.

#### Robustness check: The effect of randomizing stroke dates

Control Group 1, comprising self-reported stroke users without a known stroke date, appeared more similar to the other control groups than to the timed stroke group. This was unexpected, as both Control Group 1 and the timed stroke group consist of at-risk users. The key difference is that we lack the precise stroke date for the former.

Our hypothesis was that this discrepancy stems from how we defined a pseudo-stroke date for Control Group 1. Specifically, we chose the midpoint between each user’s first and last post and treated that as their stroke date for all time-based calculations. We suspected that this arbitrary choice could introduce noise and obscure the expected trends.

To test this, we created a new group based on the timed stroke users, but with randomized stroke dates. For each user, we selected a uniformly distributed random offset (range: plus or minus 1 year) before or after the stroke date and shifted their actual stroke date by that amount. We then recomputed the weekly attribute means using these randomized dates and compared the trends to the original timed stroke and control groups.

As an example, consider the **root TTR** attribute. Figure 3 shows the new control group in comparison to the previous 4 groups. The new group deviates from the original timed stroke group and appears closer to the control groups.

**Figure 3:**
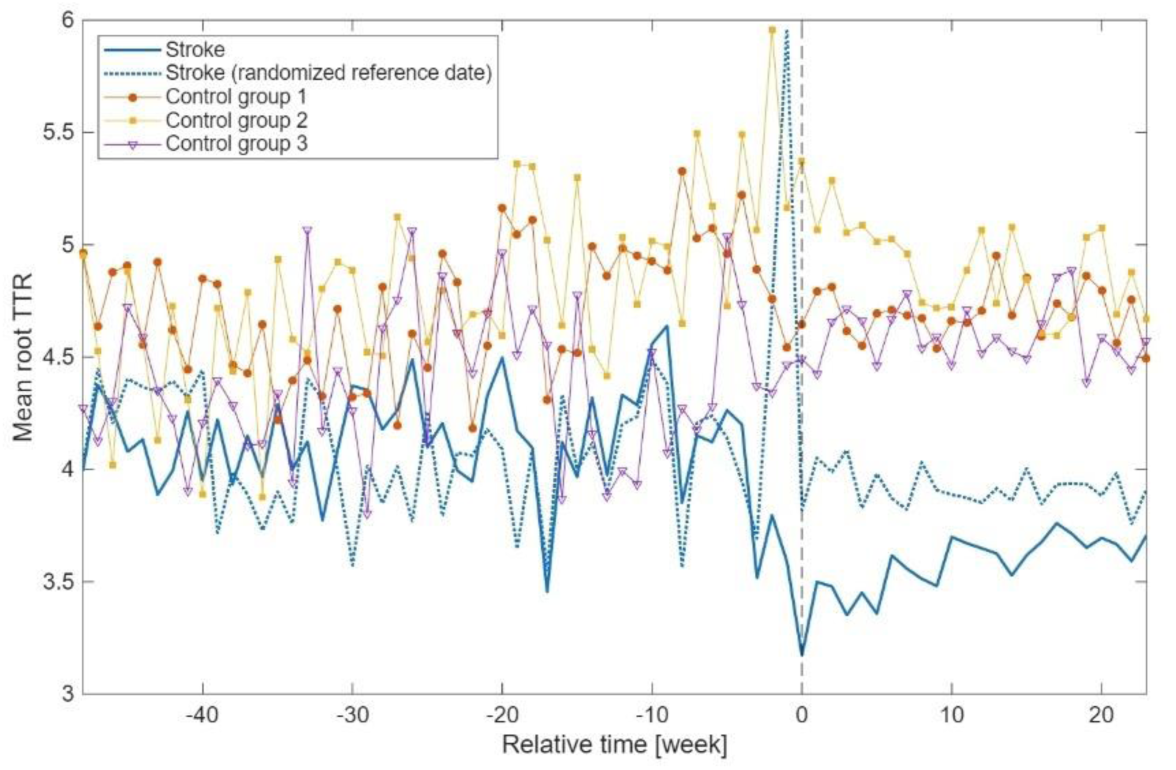
Weekly Mean Root TTR after randomizing the stroke date of the timed stroke group within a 12-month window around the stroke date. The figure shows the mean root TTR values for the stroke group, the stroke group after randomization of the reference date

Similar effects were observed for other variables. Thus, this robustness check supports our hypothesis that the difference of Control Group 1 from the timed stroke group stems primarily from the lack of knowledge on the exact stroke date. Assigning the midpoint between the first and last posts as a proxy stroke date likely introduces a large temporal error—often exceeding 4 years. Since even a 2-year offset blurs the signal substantially, it stands to reason that larger offsets would have an even greater effect.

#### Attributes Over Time - Additional variables

Figure 4 shows additional attributes over time, stratified by user group.

**Figure 4:**
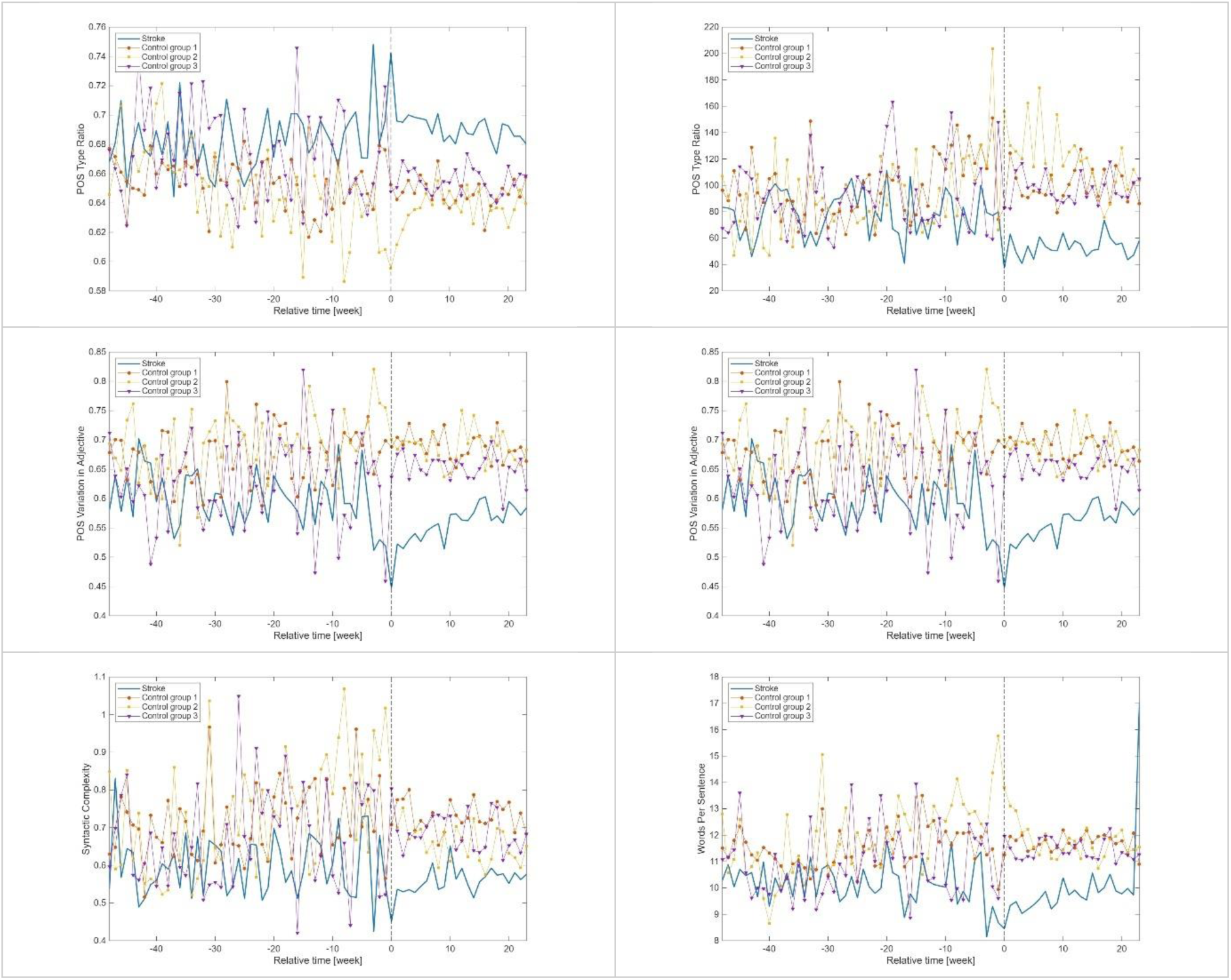
Additional attributes over time, stratified by group.

#### Comparison of Blood Pressure Mentions to Global Norms

To evaluate whether users who reported blood pressure (BP) measurements on Reddit around the time of stroke differed from the general population, we conducted a comparative analysis using real-world reference data. Specifically, we divided the timed stroke group into three time windows relative to the stroke event:

1. Before 3 Months: More than 3 months prior to the reference date.
2. Around Stroke: From 3 months before the reference date to 3 months after.
3. After 3 Months: More than 3 months after the reference date.

For each user in the timed stroke group, we analyzed all posts to identify mentions of BP readings. We searched for key terms and phrases such as “blood pressure”, “bp”, “systolic”, “diastolic”, “pressure was”, “reading”, “normal pressure”, “elevated bp”, and “low pressure”. Whenever a post contained one of these expressions, we attempted to extract the systolic (SBP) and diastolic (DBP) values using the following regular expressions:

1. Patterns similar to “120/80”: \b\d{2,3}\s*/\s*\d{2,3}\b
2. Patterns similar to “systolic 120 and diastolic 80”: systolic[^0-9]{0,5}(\d{2,3})[^a-z0-9]{1,10}diastolic[^0-9]{0,5}(\d{2,3})
3. Patterns similar to “blood pressure 140 over 90”: \bblood pressure[^0-9]{0,5}(\d{2,3})[^a-z0-9]{0,10}over[^0-9]{0,5}(\d{2,3})
4. Patterns similar to “bp 130 / 80”: \bbp[^0-9]{0,5}(\d{2,3})\s*/\s*(\d{2,3})\b
5. Patterns similar to “BP was 120”: \bbp[^a-z0-9]{0,5}(\d{2,3})\b
6. Patterns similar to “blood pressure 120”: \bblood pressure[^0-9]{0,10}(\d{2,3})\b
7. Patterns similar to “130 over 90”: \b(\d{2,3})\sover\s(\d{2,3})\b
8. Patterns similar to “135-85”: \b\d{2,3}\s*-\s*\d{2,3}\b
9. Patterns similar to “BP reading was 110/70”: bp[^a-z0-9]{0,10}reading[^0-9]{0,10}(\d{2,3}\s*/\s*\d{2,3})

The results from these expressions also included some surrounding context (text before and after the match), which facilitated manual filtering and validation of relevant entries.

After extracting these values, we computed the average SBP and DBP for each time window and compared them against global reference values.

We used the global age-standardized blood pressure statistics(27) as reference values. The global means in 2015 127/79 (men) and 122/77 (females). Since our dataset includes a mixed-gender population, we calculated a weighted average based on the proportion of males and females observed in our Reddit cohort (53% men, 47% women), which is 125/78.

For each group and BP measure (SBP and DBP), we conducted a two-tailed z-test to assess whether the sample mean significantly deviated from the global population mean.

The results are shown in Table 7. We observe that in all three time windows SBP was significantly higher than the global average, suggesting that individuals in the timed stroke group exhibited elevated systolic blood pressure across all periods, even months before and after the stroke. In contrast, DBP was significantly elevated only around the time of stroke and after it, but not in the months leading up to the event. This may indicate that DBP increases closer to the stroke event or reflects post-stroke complications or stress responses.

**Table 7:** Average blood pressure values extracted from Reddit posts in the stroke group compared to weighted average global values. SBP and DBP denote systolic and diastolic blood pressure, respectively. P-values (in bold, statistically significant P<0.05 with Bonferroni correction) denote are of the comparison against the weighted average global values.

|  |  | SBP |  | DBP |  |
| --- | --- | --- | --- | --- | --- |
| Time | N | Average | p-value | Average | p-value |
| 3 months prior | 47 | 146 | **<0.001** | 84 | 0.093 |
| Around stroke | 24 | 141 | 0.021 | 95 | 0.014 |
| 3 months post | 46 | 152 | **<0.001** | 82 | **<0.001** |

Moreover, SBP is elevated prior to stroke (146 mmHg), dips slightly around the stroke event (141 mmHg), but increases to the highest levels after 3 months (152 mmHg). This may reflect initial medical intervention followed by insufficient long-term BP control. DBP shows a marked increase around stroke (95 mmHg) compared to before (84 mmHg), and remains elevated post-stroke (92 mmHg), indicating possible lasting physiological effects of the stroke or ongoing stress responses. These patterns highlight the importance of sustained post-stroke monitoring and intervention for blood pressure management.

Overall, the findings highlight a pattern of persistent systolic hypertension among users in the stroke cohort, which may be a contributing risk factor or an indicator of insufficient BP management post-stroke.

